# Trends in transitions in cigarette and electronic nicotine vapor product use in the Rutgers Omnibus Study, 2022–2026

**DOI:** 10.64898/2026.09.28.26364207

**Authors:** Olivia K. Roberts, Michelle T. Bover Manderski, Nishi J. Gonsalves, Jihyoun Jeon, Rafael Meza, Cristine D. Delnevo, Andrew F. Brouwer

## Abstract

**Introduction:** Monitoring transitions in cigarette and electronic nicotine vapor product (ENVP) use is important for understanding how marketplace and regulatory changes have affected product use patterns, but information from national surveys is often delayed by several years.

**Methods:** The Rutgers Omnibus Study is a quarterly serial, rapid-response, web-based convenience survey. Using a Markov multistate transition model, we estimated transition rates and one-year transition probabilities between never, non-current, ENVP-only, cigarette-only, and dual use among 5,292 adults aged 18–45 who participated in at least two of Waves 1–16 (February 2022–February 2026), as well as overall transition hazard ratios by sex, age group, and race & ethnicity in a multivariable model.

**Results:** About 40% of individuals using cigarettes (39.5%; 95% CI: 36.9–42.0) or ENVP (40.1%; 95% CI: 36.4–43.7), and 60% of those using both products (60.8%; 95% CI: 58.3–63.2) were predicted to remain in those states after one year. Female participants began using cigarettes from never use (adjusted hazard ratio [aHR] 0.44; 95% CI: 0.29–0.66) or non-current use (aHR 0.65; 95% CI: 0.48–0.88) at lower rates than male participants. Participants aged 35–45 years initiated ENVP-only use from non-current use (aHR 0.34; 95% CI: 0.23–0.51) at a lower rate than those aged 18–34 years. Compared to non-Hispanic White participants, non-Hispanic Black participants initiated ENVP-only use from never use at a higher rate (aHR 4.39; 95% CI: 2.09–9.25).

**Conclusion:** Rapid response surveys can provide important preliminary information on tobacco use transition patterns while we wait for the slower release of nationally representative survey data.

**What this paper adds:** *What is already known on this topic:*

- The use of electronic nicotine vapor products (ENVPs) has continued to evolve over time—as well as attitudes towards them—but the release of data from large nationally representative surveys often lags behind these changes.

*What this study adds:*

- The Rutgers Omnibus Study provides rapid response data on tobacco use trends.
- Transition probabilities between tobacco use states remained relatively stable over time. Participants in this sample frequently switched between tobacco products, but those using both cigarettes and ENVPs were very likely to continue using one or both products.
- Female participants began using cigarettes at lower rates than male participants, participants ages 35–45 years began using ENVPs at a lower rate than participants ages 18–34 years, and non-Hispanic Black participants initiated use of ENVPs at a higher rate than non-Hispanic White participants.

*How this study might affect research, practice or policy:*

- Rapid response tobacco use surveys can help fill the gap in product use trends and transitions while we await the slower release of nationally representative data, such as the Population Assessment of Tobacco and Health (PATH) Study.

## Introduction

Electronic nicotine vapor products (ENVPs) have continued to evolve in recent years, alongside a shifting retail environment. For example, from 2022 to 2023, overall unit sales of e-cigarettes declined by 4.8%, yet there was an increase in total nicotine content sold by 15.5%, suggesting a transition towards higher-concentration nicotine products.[1] At the same time, attitudes towards ENVPs have evolved; a growing majority of adults incorrectly believe that e-cigarettes are equally or more harmful than combustible cigarettes.[2] Tobacco control efforts often lag behind changes in the tobacco marketplace, and new products can remain on the market for years before regulations are implemented.[3] Lags in data collection and availability contribute to lags in regulation, so timely data on ENVP and cigarette use trends are essential to address emerging risks and inform regulations as the marketplace continues to evolve.

Multistate transition models have been applied to nationally representative longitudinal studies such as the Population Assessment of Tobacco and Health (PATH) Study to estimate underlying transition rates between tobacco use states over time and by sociodemographic characteristics.[4,5] However, larger longitudinal studies such as PATH, which are invaluable to the field, also necessarily release data more slowly because of the infrastructure involved (at the time of writing in 2026, PATH Wave 8 public-use data from 2024 have just been released). Rapid-response tobacco use surveillance surveys can fill the gap while we await data from larger nationally-representative studies.[6] This analysis examines adult product use transitions in the rapid-response Rutgers Omnibus Study over time and by sociodemographic group from February 2022 to February 2026. The multistate transition modeling approach allows us to estimate the underlying transition rates that lead to observed patterns of cigarette and ENVP use and identify trends in these transitions over time.

## Methods

### Data

The Rutgers Omnibus Study, a rapid-response, convenience-sample survey conducted quarterly beginning in February 2022, provides rapid data collection through online Amazon Mechanical Turk surveys to monitor the use of tobacco and nicotine products among adults aged 18–45 in the United States.[7] Participants were recruited through the CloudResearch Mturk Toolkit for improved data quality over direct Mturk recruitment.[8,9] The Omnibus study was approved as exempt by the Rutgers University Institutional Review Board as the data were collected in such as a way that participant’s identities cannot be readily identified.

Although the Rutgers Omnibus Study was originally intended as a repeated cross-sectional study rather than a longitudinal panel study, individuals were not excluded from responding to multiple surveys, and 6,459 individuals had two or more observations across collection Waves 1–16, providing 24,385 total longitudinal observations. For this analysis, participants with missing responses to questions about cigarette or ENVP use were excluded, as were those with missing or inconsistent answers to questions about sex, race & ethnicity, and age. For each wave, participants were classified into tobacco use states based on established and past 30-day use of each product. Consistent with previous work using PATH data,[4,5] we defined five product use states: never use (of either product), non-current use (of either product), cigarette-only use, ENVP-only use, and dual use of cigarettes and ENVP. Established use of cigarettes was defined as use of 100+ lifetime cigarettes, while established use of ENVP was defined as any previous use. Current use of a product was defined as use on at least one day in the past 30 days, while non-current use was defined as established use without any past 30-day use. If an individual answered “yes” to ever use of cigarettes, ever use of ENVP, or 100+ lifetime cigarettes use but “no” in their next observation, that subsequent observation was removed due to inconsistency. The final sample used for the analysis across all Waves contained 18,142 observations of 5,292 individuals, with at least two observations per individual. In total, about 26% of the original observations were removed. While such a large percentage removal would be a concern in a nationally representative study, this degree of data cleaning is not unexpected for a web-based convenience sample. The data collection date for responses in each Wave was set to the midpoint of the survey’s open period.

To track changes in transition probabilities over time, we grouped the survey waves into four time periods of approximately one year: Waves 1–5 (February 2022 to February 2023; abbreviated as 2022), 5–8 (February 2023 to January 2024; abbreviated as 2023), 8–12 (January 2024 to February 2025, abbreviated as 2024), and 12–16 (February 2025 to February 2026; abbreviated as 2025). Periods were defined with overlapping boundaries to ensure that transitions across boundary waves were included. Within each of the four periods, only individuals with at least two observations in that period were included. The four periods had 5,186; 4,450; 3,488; and 2,753 observations of 2,131; 1,757; 1,439; and 1,336 individuals, respectively. Prevalence estimates for cigarette-only use, ENVP-only use, and dual use, as well as characteristics of the population by sex, age group, and race & ethnicity, were determined using each participant’s first observation within each period (see Table S1 in the Supplementary Material). In addition to estimating transition probabilities over time, we calculated the p-values for trends in prevalence and transition probabilities across these periods using simple linear regression, assuming equal one-year intervals between periods.

### Transition modeling

We use a Markov multistate transition model to estimate the underlying transition hazard rates between product use states for adults overall, by covariate (sex, age group, and race & ethnicity), and by time period. A Markov multistate transition model assumes that transitions arise from a continuous-time, finite-state stochastic process in which transition rates depend only on the current state, not on past states or transition history (the Markov property). The model estimates transition hazard rates, which are the instantaneous risks of transitioning from one state to another. Technical details of multistate modeling are provided in the Supplementary Material. State definitions and allowed transitions are given in Figure S1 in the Supplementary Material. Details of the analysis used to determine the allowed vs negligible transitions in the model are given in the Supplementary Material. Transition rates were estimated using the msm function from the msm package in R (v1.8.2).[10]

One-year transition probabilities were estimated across the entire four-year period and also estimated separately for each of the four one-year periods to assess changes over time. Transition hazard ratios by sex (female vs. male), age group (35–45 vs. 18–34), and race & ethnicity (non-Hispanic Black vs. non-Hispanic White) were calculated for the overall period using both unadjusted models and an adjusted multivariable model that included sex, age group, and race & ethnicity as covariates.

## Results

The characteristics of all participants at their enrollment Wave—including sex, age group, race and ethnicity, and educational attainment—are given in Table 1. The prevalence of cigarette-only use among participants decreased slightly overall from 14.5% in 2022 to 10.7% in 2025 (p=0.23). ENVP-only use remained virtually unchanged from 8.0% in 2022 to 8.7% in 2025 (p=0.51), while dual use increased slightly from 14.5% to 17.5% (p=0.29). However, none of these trends were statistically significant.

**Table 1.** Characteristics of the population at enrollment across all 16 waves of the Rutgers Omnibus Study (February 2022 to February 2026).

| Characteristic | Overall<br>N = 5,292 | Never use<br>N = 2,164 | Non-current<br>use<br>N = 1,166 | Cigarette-only<br>use<br>N = 755 | ENVP-only<br>use<br>N = 452 | Dual use<br>N = 755 |
| --- | --- | --- | --- | --- | --- | --- |
| Sex % (n) |  |  |  |  |  |  |
| Male | 40.6% (2,150) | 37.5% (811) | 42.1% (491) | 46.2% (349) | 36.1% (163) | 44.5% (336) |
| Female | 59.4% (3,142) | 62.5% (1,353) | 57.9% (675) | 53.8% (406) | 63.9% (289) | 55.5% (419) |
| Age group %<br>(n) |  |  |  |  |  |  |
| 18-34 | 53.2% (2,814) | 53.8% (1,164) | 49.5% (577) | 46.6% (352) | 67.5% (305) | 55.1% (416) |
| 35-45 | 46.8% (2,478) | 46.2% (1,000) | 50.5% (589) | 53.4% (403) | 32.5% (147) | 44.9% (339) |
| Race/ethnicity<br>% (n) |  |  |  |  |  |  |
| NH White | 69.9% (3,697) | 66.4% (1,436) | 72.8% (849) | 73.8% (557) | 69.0% (312) | 71.9% (543) |
| NH Black | 10.0% (527) | 12.1% (262) | 8.3% (97) | 8.2% (62) | 8.8% (40) | 8.7% (66) |
| Others | 20.2% (1,068) | 21.5% (466) | 18.9% (220) | 18.0% (136) | 22.1% (100) | 19.3% (146) |
| Educational<br>attainment %<br>(n) |  |  |  |  |  |  |
| Less than<br>high school | 1.4% (73) | 0.9% (19) | 1.3% (15) | 2.5% (19) | 0.7% (3) | 2.3% (17) |
| High school | 10.3% (546) | 7.3% (157) | 9.3% (109) | 15.4% (116) | 12.4% (56) | 14.3% (108) |
| Some<br>college or AA<br>degree | 29.2% (1,544) | 19.5% (421) | 30.5% (356) | 37.2% (281) | 37.4% (169) | 42.0% (317) |
| BA degree<br>or higher | 53.2% (2,815) | 66.0% (1,428) | 54.0% (630) | 42.4% (320) | 37.6% (170) | 35.4% (267) |
| Not<br>completed or<br>unknown | 5.9% (314) | 6.4% (139) | 4.8% (56) | 2.5% (19) | 11.9% (54) | 6.1% (46) |

Note: Educational attainment was classified as “not completed” for participants younger than 25 years. ENVP = electronic nicotine vapor product, NH = non-Hispanic, AA = associate’s degree, BA = bachelor’s degree.

### Transition probabilities for participants overall across all waves

Across all waves combined (2022–25), one-year transition probabilities indicated that the cigarette-only, ENVP-only, and dual use states were largely transient, as self-reported (Figure 1). We estimated that only 39.5% (95% CI: 36.9–42.0%) of individuals reporting smoking cigarettes only would remain in that state in one year, while 38.0% (95% CI: 35.7–40.5%) would transition to dual use and 17.4% (95% CI: 15.6–19.3%) would not be using either cigarettes or ENVPs. We estimated that, among individuals using ENVPs only, 40.1% (95% CI: 36.4–43.7%) would remain in that state, while 23.4% (95% CI: 20.6–26.3%) would transition to dual use, and 29.0% (95% CI: 25.7– 32.5%) would not use either cigarettes or ENVPs after one year. Individuals using both cigarettes and ENVPs were very likely to continue using one or both products, with 60.8% (95% CI: 58.3–63.2%) estimated remaining in the dual use state, 22.0% (95% CI: 20.0–24.0%) transitioning to cigarette use only, 10.1% (95% CI: 8.7–11.6%) transitioning to ENVP use only, and only 7.2% (95% CI: 6.5–7.9%) not using either product.

**Figure 1:**
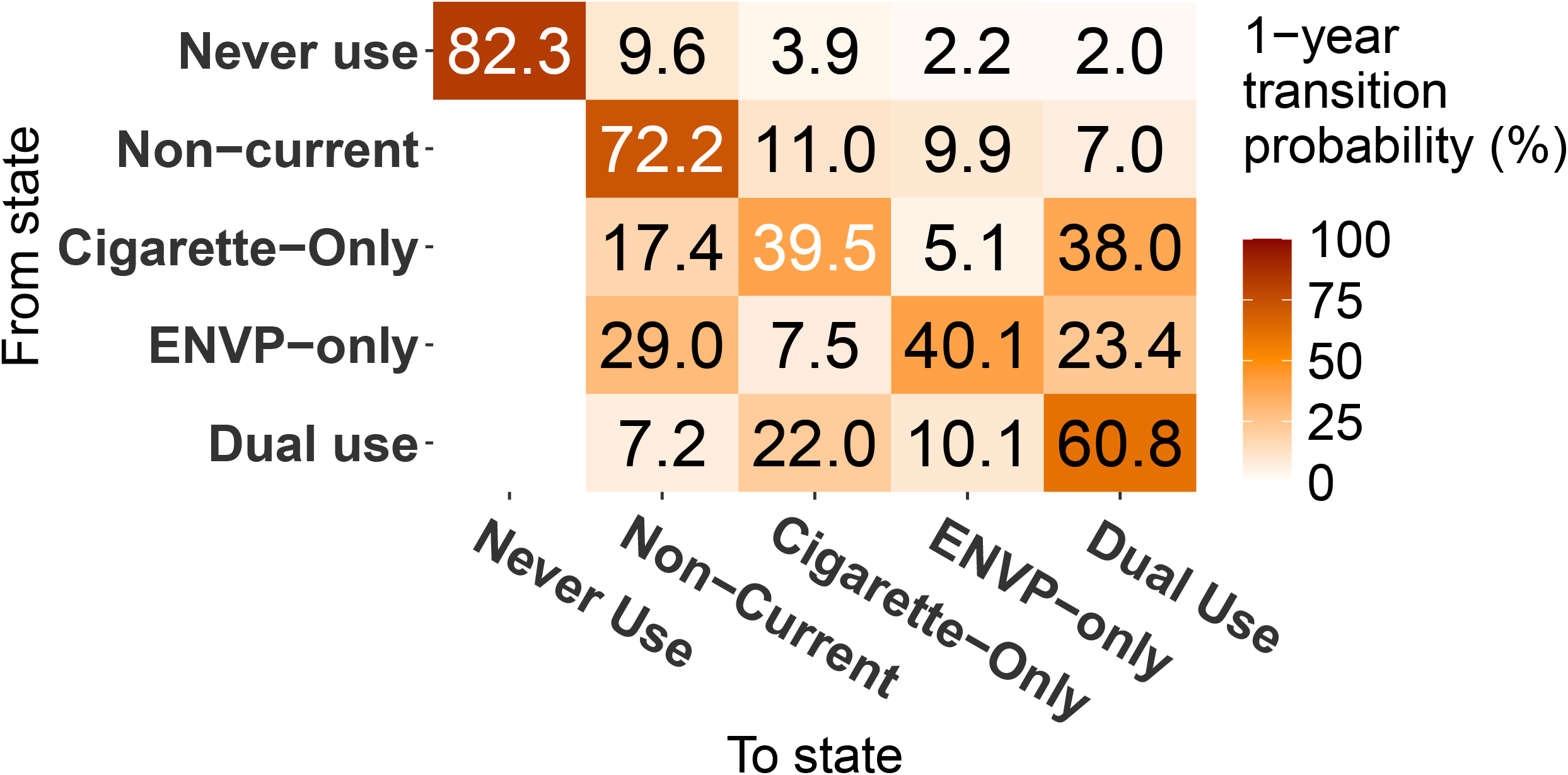
One-year transition probabilities estimated across all 16 waves of the Rutgers Omnibus Study (February 2022 to February 2026). Confidence intervals are provided in Supplementary Table S2. ENVP = electronic nicotine vapor product.

### Transition probabilities for participants overall by period

Characteristics of participants by period are given in Table S1 in the Supplementary Material. Most one-year transition probabilities remained mostly stable across the four periods (Figure 2), with no statistically significant overall trends observed for any transition over time. In 2022, about 7.1% (95% CI: 5.7–9.0%) of participants with no established use of either product transitioned to cigarette-only use, while 3.2% (95% CI: 2.4–4.6%) transitioned to ENVP-only use. By 2025, these estimates were 5.1% (95% CI: 3.6–7.4%) and 3.8% (95% CI: 2.6–6.2%), respectively, with no significant trends over time (p=0.32 for never to cigarette-only use, p=0.54 for never to ENVP-only use). Other transitions without significant trends over time were non-current to cigarette-only use (p=0.06), non-current to ENVP-only use (p=0.21), cigarette-only to non-current use (p=0.95), ENVP-only to non-current use (p=0.58), ENVP-only to dual use (p=0.99), and dual to ENVP-only use (p=0.91).

**Figure 2:**
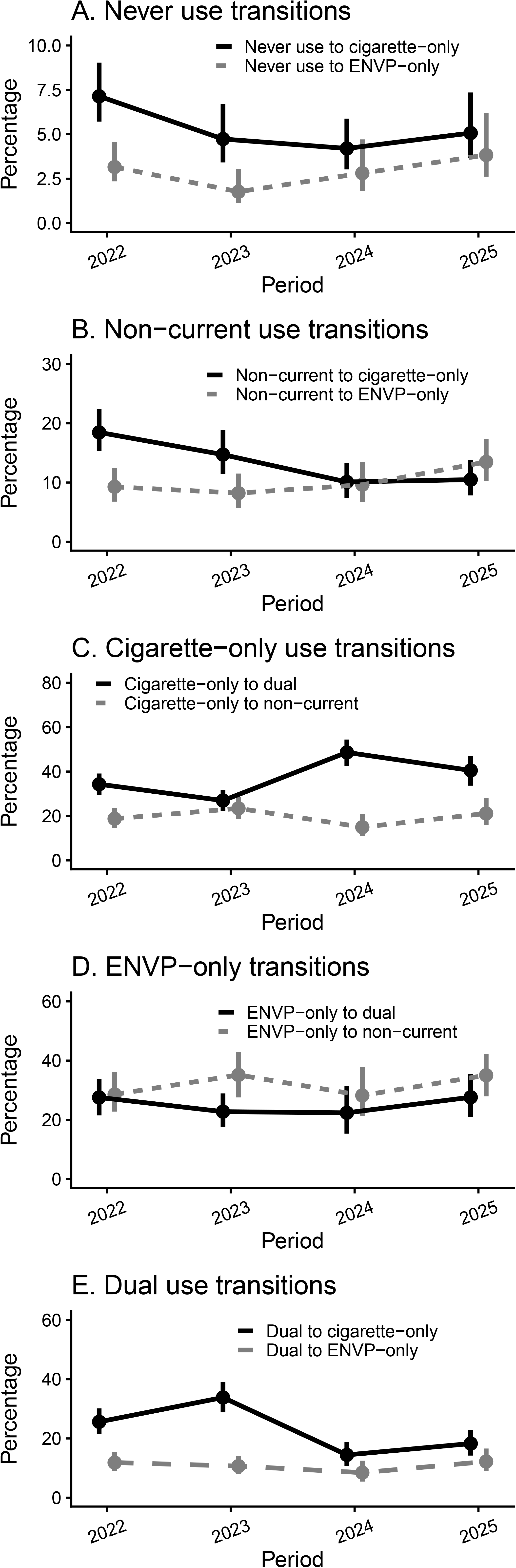
One-year transition probabilities and 95% confidence intervals for a) never use, b) non-current use, c) cigarette-only use, d) Electronic nicotine vapor product (ENVP)-only use, and e) dual use, estimated by period of the Rutgers Omnibus Study: Waves 1–5 (February 2022 to February 2023; abbreviated as 2022), 5–8 (February 2023 to January 2024; abbreviated as 2023), 8–12 (January 2024 to February 2025, abbreviated as 2024), and 12–16 (February 2025 to February 2026; abbreviated as 2025). All point estimates and confidence intervals are provided in Supplementary Table S2.

There were some statistically significant changes in one-year transition probabilities from 2023 to 2024 only; transitions from cigarette-only use to dual use increased from 26.9% (95% CI: 22.2–31.8%) in 2023 to 48.6% (95% CI: 42.4–54.4%) in 2024. During this same period, there was an analogous decrease in transitions from dual use to cigarette-only use from 33.8% (95% CI: 28.9–39.1%) to 14.4% (95% CI: 10.7–18.8%). However, there were no significant trends for these transitions across the entire period overall (p=0.44 for cigarette-only to dual use, p=0.38 for dual to cigarette-only use).

### Transition hazards by sex, age group, and race & ethnicity

Over the entire period, female participants initiated or relapsed to cigarette use at a lower rate than male participants (Figure 3). From never use, female participants began using cigarettes at about half the rate as male participants (adjusted hazard ratio [aHR] 0.44; 95% CI: 0.29–0.66). Female participants also transitioned with a lower rate from non-current to cigarette-only use (aHR 0.65; 95% CI: 0.48–0.88), ENVP-only to dual use (aHR 0.54; 95% CI: 0.37–0.80), and cigarette-only to non-current use (aHR 0.59; 95% CI: 0.42–0.81).

**Figure 3:**
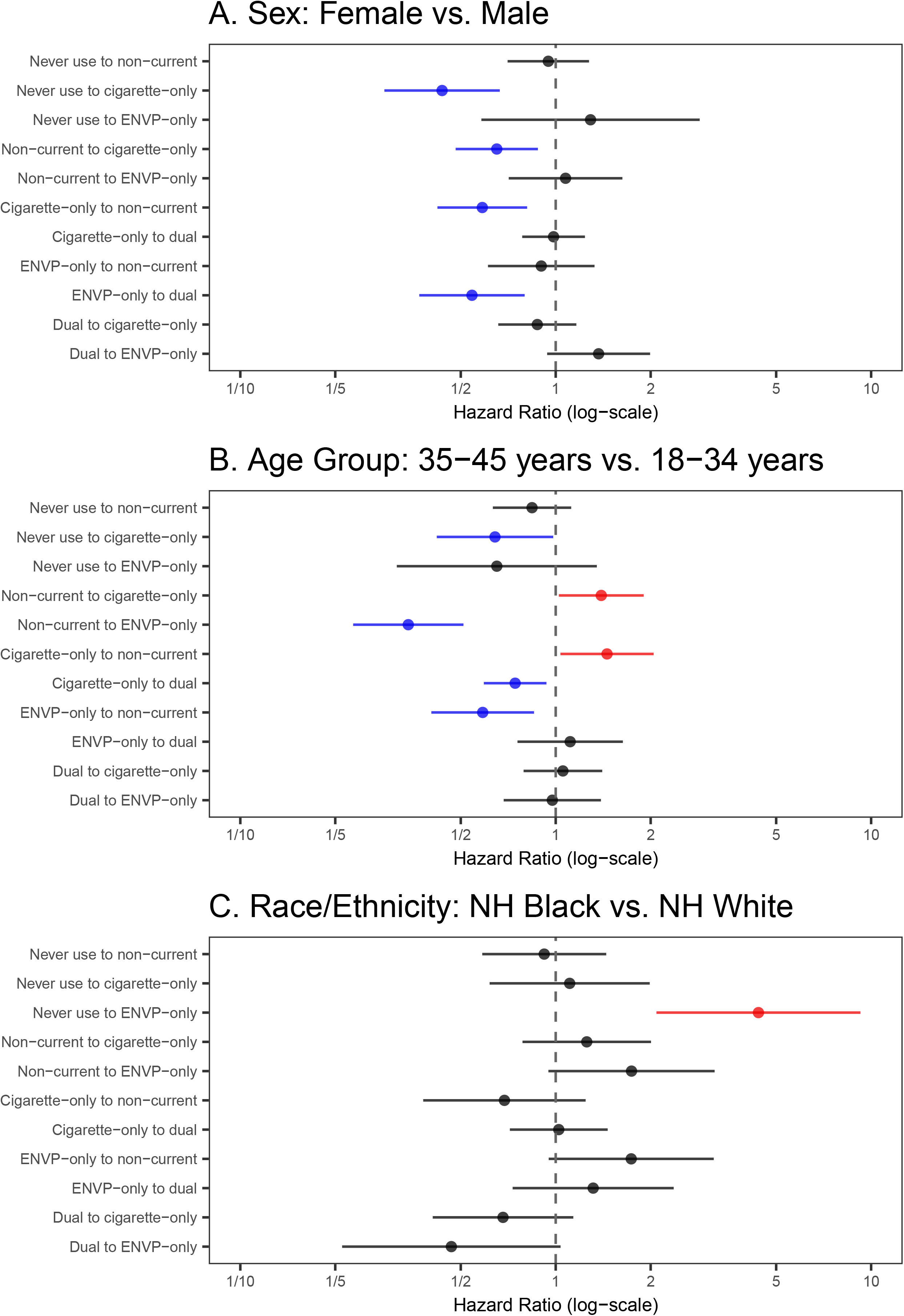
Transition hazard ratios estimated across all 16 waves of the Rutgers Omnibus Study (February 2022 to February 2026) for a) sex, b) age group, and c) race & ethnicity in multivariable models adjusted by sex, age group, and race & ethnicity. Statistically significantly greater hazard ratios are in red, and statistically significantly lower hazard ratios are in blue. All point estimates and confidence intervals are provided in Supplementary Table S3. ENVP = electronic nicotine vapor product.

There were significant differences in transition rates by age group. Compared to those ages 18–34 years, participants ages 35–45 years transitioned from never to cigarette-only use (smoking initiation) at a lower rate (aHR 0.64; 95% CI: 0.42–0.98) but from non-current to cigarette-only use (smoking relapse) at a higher rate (aHR 1.39; 95% CI: 1.02–1.90) and cigarette-only to non-current use (smoking discontinuation) at a higher rate (aHR 1.45; 95% CI: 1.03–2.04). The older age group transitioned from non-current to ENVP-only use (aHR 0.34; 95% CI: 0.23–0.51) and cigarette-only to dual use (aHR 0.74; 95% CI: 0.59–0.93) at lower rates than the younger age group. The older age group also transitioned from ENVP-only to non-current use at a lower rate (aHR 0.59; 95% CI: 0.40–0.85). Compared to non-Hispanic White participants, non-Hispanic Black participants initiated ENVP-only use from never use at a higher rate (aHR 4.39; 95% CI: 2.09–9.25).

All estimated one-year transition probabilities overall and by period with confidence intervals are provided in Table S2 in the Supplementary Material. All unadjusted and adjusted hazard ratios by covariate with confidence intervals are provided in Table S3 in the Supplementary Material.

## Discussion

We estimated transition rates for the initiation and cessation of cigarette and ENVP products in the quarterly Rutgers Omnibus Study from February 2022 to February 2026 and transition hazard ratios by sex, age group, and race & ethnicity. In this convenience sample, individuals reported frequently switching between tobacco products, discontinuation, and relapse. However, individuals using both products were very likely to continue using one or both products. There was relatively low persistence in any single product use state, and these transition probabilities remained largely stable over time. Female participants began using cigarettes at lower rates than male participants in this sample, including transitions from never use to cigarette-only use, non-current use to cigarette-only use, and ENVP-only use to dual use. Participants ages 35–45 years began using ENVP products from non-current or cigarette-only use at a lower rate than participants ages 18–34 years. While the older age group transitioned from never use to cigarette-only use at a lower rate than the younger age group, they transitioned from non-current to cigarette-only use at a higher rate. Compared to non-Hispanic White participants, non-Hispanic Black participants initiated use of ENVPs from never use at a higher rate.

One advantage of this analysis is that—at this time—there are no nationally representative longitudinal datasets that include 2026. The most recent comparable analysis by Brouwer et al. using nationally representative PATH data reports hazard ratios and one-year transition probabilities in 2021–24 compared to previous years.[5] A limitation of using recent rapid-response data is that direct numeric comparisons with nationally representative studies are challenging, but qualitative comparisons can provide insights into the reliability of our estimates. Compared to the 2021–24 PATH analysis, transition probabilities in this analysis differ in magnitude, with higher probabilities of initiation or product switching. In this analysis, among individuals who have never used either product, 4.0% initiate cigarette-only use, 2.2% initiate ENVP-only use, and 2.0% initiate dual use in one year, while the most recent PATH estimates for 2022–24 (Waves 7–8) for these transitions were considerably lower at 0.1%, 0.3%, and 0.0%, respectively. Individuals in this analysis using only cigarettes or ENVPs were more likely to transition to dual use compared to the PATH analysis; 38.0% of those using only cigarettes and 23.4% of those using only ENVPs transitioned to dual use, compared to 5.9% and 4.5% in PATH. Individuals in this analysis using only one product were less likely to remain in the same state after one year compared to PATH (39.5% compared to 86.7% for cigarettes, and 40.1% compared to 79.4% for ENVPs), while individuals using both products were about as likely to continue using both products after one year compared to PATH (60.8% compared to 62.3%). The more transient nature of product use in the Rutgers Omnibus Survey may reflect differences in the frequency of data collection. If PATH were measured quarterly rather than annually or biennially, we might find evidence of more frequent transitions. Also, there were differences in educational attainment between the two datasets. The Rutgers Omnibus Survey sample had a high level of educational attainment; over half of participants had a bachelor’s degree or higher (Table 1), compared with approximately a third of participants on average across PATH survey Waves.[4] Differences in educational attainment may have contributed to differences in product use transitions between the two datasets, although the extent of this relationship is unclear.

Despite differences in magnitude, many patterns remain similar between these two datasets. In both PATH and Rutgers Omnibus Survey analyses, the probability of transitioning from non-current use to cigarette-only, ENVP-only, or dual use is higher than the probability of transitioning from never use to each product use state, respectively. Also, in both analyses, the probability of remaining in the same state after one year is higher than any individual probability of transitioning to a different use state. Some transition hazard ratios by age group were also qualitatively similar. In both analyses, older age groups transitioned from non-current use to cigarette use at a higher rate, while younger age groups transitioned from non-current to ENVP and cigarette to dual use at higher rates.

Multistate transition modeling is becoming more commonly utilized in tobacco control to investigate transition rates between tobacco use states in longitudinal studies.[11,12] However, this method and our study have some limitations. The Markov property of multistate transition models assumes that transition probabilities do not depend on past states or duration of past states, which means that individuals’ full history of initiation or cessation was directly considered. Therefore, a caution in interpreting this analysis is that our results capture population averages, not individual trajectories. Moreover, as a convenience sample, the Rutgers Omnibus Study is not nationally representative, so these results cannot be generalized to the broader U.S. population. The use of Mechanical Turk (MTurk) to collect data enables rapid data collection, but there remain concerns about the reliability of MTurk data.[13] To address these concerns, the

Rutgers Omnibus Study recruits only pre-approved MTurk workers from the CloudResearch MTurk Toolkit,[9] and this analysis implemented data quality control measures by excluding participants with missing responses and inconsistent demographic information (sex, age, and race & ethnicity) and removing observations with impossible transitions. The relative stability of transition probabilities over time in this analysis suggests that there is consistency within the sample, even if it is not nationally generalizable.

This analysis does not directly account for oral nicotine pouch (ONP) use, which started to increase in the overall U.S. population between 2023 and 2024.[14] Indeed, the prevalence of ONP use among participants in the final sample used for this analysis increased over the period from approximately 0.5% in Wave 1 to a bit less than 5% in Wave 16. However, the prevalence in each Wave remained low overall (less than 5%), and as discussed, there were few significant changes in cigarette and ENVP transitions during this period. It is possible that, counterfactually, without ONP use, we would have seen some meaningful trends, but it is more likely that the prevalence of ONP use was too low at this point to have any influence. Fortunately, the rapid response approach is likely to detect and report changes much earlier than will be possible in PATH or other large surveys. The use of ONP and ENVP in this survey is continually monitored and will be evaluated in future work.

## Conclusion

Participants in the Rutgers Omnibus Study frequently switched between cigarette and ENVP product use patterns; however, those using both cigarettes and ENVPs were very likely to continue using one or both products, highlighting the importance of tobacco control efforts aimed at reducing all tobacco product initiation. Differences by sociodemographic group highlight disparities in use patterns and transitions. Continued monitoring of these transitions is essential to identify any emerging changes in tobacco product use over time. While we wait for the slower release of nationally representative survey data from longitudinal studies such as PATH, rapid response surveys can provide important preliminary information on tobacco use transition patterns to inform tobacco control efforts in a rapidly evolving product marketplace.

## Acknowledgements

This project was funded through National Cancer Institute (NCI) and Food and Drug Administration (FDA) grants U54CA229974 and U01CA278695. The opinions expressed in this article are the authors’ own and do not necessarily reflect the views of the National Institutes of Health, the Department of Health and Human Services, or the United States government.

## Contributors

Conceptualization: CDD, MTB-M, AFB, RMe; methodology: AFB; data curation: MTB-M, NJG, OKR; analysis: OKR; original draft preparation: OKR; review and editing: MTB-M, NJG, JJ, RMe, CDD and AFB; funding acquisition: CDD, MTB-M, RMe; guarantor: AFB.

## Data Availability Statement

Study measures are available upon request to authors. Data from Waves 1–16 of the Rutgers Omnibus Survey are not publicly available in a repository because the 2002–2005 online consent did not indicate that the data could be placed in a repository. A restricted dataset may be requested from PI of the Rutgers Omnibus Survey, Michelle Bover Manderski, and should include a plan for its use. All data sharing will comply with local, state, and federal laws and regulations and may be subject to appropriate human subjects institutional review board approvals.

