## Supplementary material for "Trends in transitions in cigarette and electronic nicotine vapor product use in the Rutgers Omnibus Study, 2022–2026"

**Multistate transition modeling**

This technical appendix is reproduced in part from the supplementary material of Brouwer et al [4] but includes updated notation and technical details. Additional details may be found in Jackson [7].

A multistate transition model is a continuous-time, finite-state stochastic process with the Markov assumption that transition rates depend only on the current state and not on past states or transition history. We denote the state of the process at time $t$ as $S\left( t \right)$. We denote the probability that an individual is in state $j$ after an amount of $\Delta t$ since they were observed in state $i$ as

$$\begin{aligned} P_{ij}(t,t+\Delta t)=Pr\left[ S\left( t+\Delta t \right)=j | S\left( t \right)=i \right]. \#\left( 1 \right) \end{aligned}$$

In general, the transition probabilities can depend on the observation time $t$ in addition to the time span $\Delta t$, and in this analysis we consider discrete time periods over which we assume that that the model is homogeneous in time, i.e., $P_{ij}\left( t,t+\Delta t \right)= P_{ij}(0,\Delta t)$. Thus, we drop the dependence on $t$ moving forward and write $P_{ij}(\Delta t)$. We then define the hazard rate of the transition from state $i$ to state$j$, for $i\neq j$, as

$$\begin{aligned} q_{ij}=\lim_{\Delta t\to0} \frac{1}{\Delta t}Pr\left[ S\left( \Delta t \right)=j | S\left( 0 \right)=i \right]. \#\left( 2 \right) \end{aligned}$$

Transition hazards ratios $\rho_{ij,c_{l}}$may be determined for each transition $i$ to $j$ for each level $l$ of a covariate $c$, relative to the referent $c_{0}$: $q_{ij,c_{l}}=\rho_{ij,c_{l}}\times q_{ij,c_{0}}$. For time-varying covariates, a participant’s transition rate is determined by their characteristic at their most recent previous observation.

The transition hazard rates form a matrix $Q = \left[ q_{ij} \right],$ where the diagonal entries are given by $q_{ii}=-\sum_{j\neq i} q_{ij}$. The transition probability matrix $P\left( \Delta t \right)=\left[ P_{ij}\left( \Delta t \right) \right]$ is a function of the transition hazard rates and may be calculated as the matrix exponential of $\Delta t\cdot Q$, that is

$\begin{aligned} P\left( \Delta t \right)=\text{expm}\left( \Delta t\cdot Q \right).\#\left( 3 \right) \end{aligned}$

Transition probabilities can be estimates for any value of $\Delta t$, assuming that the transition hazards $q_{ij}$ do not change over that time period (assumption of homogeneity).

The values of the transition hazard rates $q_{ij}$of multistate transition model are estimated by maximizing a statistical likelihood $L$ given a set of observed states and times by comparing the observed states to the probabilities in $P\left( \Delta t \right)$ as a function of the transition hazard matrix $Q$. Specifically, consider a set of individuals $m = 1, \ldots,N$ and their observed states $s_{m,t_{m,k}}$at times $t_{m,k}$, where $k$ is the index of individual $m$’s $K_{m}$observations in the data. Denote the data as $s=\{s_{m,t_{m,k}}\}$. We assume the individuals are independent, and thus we multiply all the modeled probabilities of the observed transitions:

$$\begin{aligned} L\left( Q|s \right)=\prod_{m=1}^{N} \prod_{k=1}^{K_{m}-1} P_{s_{m,t_{m,k}},s_{m,t_{m,k+1}}}(t_{m,k+1}-t_{m,k}).\#\left( 4 \right) \end{aligned}$$

where $P_{i,j}$ is a function of $Q$ as in Eqn (3).

The set of direct transitions allowed between states in the model was first restricted to transitions that were theoretically plausible. Among these, the final set of allowed transitions was selected by minimizing a Bayesian Information Criterion (BIC), given by

$$\begin{aligned} BIC=p \text{log(}n\text{)-2}\cdot\text{log}L\#\left( 5 \right) \end{aligned}$$

Where $p$ is the number of parameters, $n$ is the number of observations, and $L$ is the maximized likelihood of the model.

***Figure S1****. a) State definitions and b) allowed instantaneous transitions in the model.*

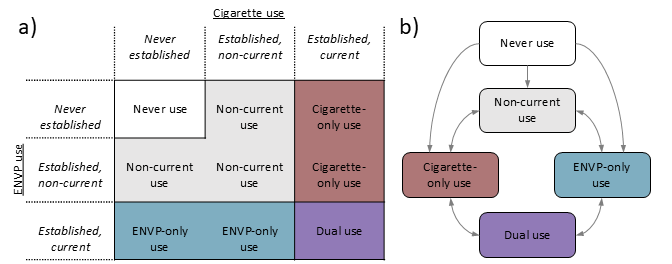

***Table S1****: Characteristics of participants in the Rutgers Omnibus study by period: Waves 1–5 (February 2022 to February 2023; abbreviated as 2022), 5–8 (February 2023 to January 2024; abbreviated as 2023), 8–12 (January 2024 to February 2025, abbreviated as 2024), and 12–16 (February 2025 to February 2026; abbreviated as 2025). Note that in this table, characteristics are listed for each unique individual’s first observation within the period.*

| **Characteristic** | **2022 (Waves 1–5)**  N = 2,131 | **2023 (Waves 5–8)**  N = 1,757 | **2024 (Waves 8–12)**  N = 1,439 | **2025 (Waves 12–16)**  N = 1,336 |
| --- | --- | --- | --- | --- |
| Sex, % (n) |  |  |  |  |
| Male | 42.3% (901) | 40.5% (711) | 43.3% (623) | 43.6% (582) |
| Female | 57.7% (1,230) | 59.5% (1,046) | 56.7% (816) | 56.4% (754) |
| Age group, % (n) |  |  |  |  |
| 18-34 | 52.6% (1,121) | 50.1% (881) | 44.1% (634) | 40.9% (547) |
| 35-45 | 47.4% (1,010) | 49.9% (876) | 55.9% (805) | 59.1% (789) |
| Race/ethnicity, % (n) |  |  |  |  |
| Non-Hispanic White | 70.3% (1,498) | 68.7% (1,207) | 70.3% (1,011) | 70.9% (947) |
| Non-Hispanic Black | 9.5% (203) | 10.4% (182) | 10.5% (151) | 9.2% (123) |
| Others | 20.2% (430) | 20.9% (368) | 19.2% (277) | 19.9% (266) |
| Use status, % (n) |  |  |  |  |
| Never use | 41.7% (889) | 40.5% (711) | 41.5% (597) | 38.1% (509) |
| Non-current use | 21.4% (455) | 21.8% (383) | 22.2% (319) | 25.0% (334) |
| Cigarette-only use | 14.5% (308) | 14.7% (258) | 14.5% (208) | 10.7% (143) |
| ENVP-only use | 8.0% (170) | 7.6% (133) | 7.5% (108) | 8.7% (116) |
| Dual use | 14.5% (309) | 15.5% (272) | 14.4% (207) | 17.5% (234) |
| Note: Other race/ethnicity includes Hispanic participants and non-Hispanic participants of other and multiple races. | | | | |

***Figure S2****. Unadjusted transition hazard ratios estimated across all 14 waves of the Rutgers Omnibus study (February 2022 to February 2026) for a) sex, b) age group, and c) race & ethnicity in univariable models. Statistically significantly greater hazard ratios are in red, and statistically significantly lower hazard ratios are in blue.*

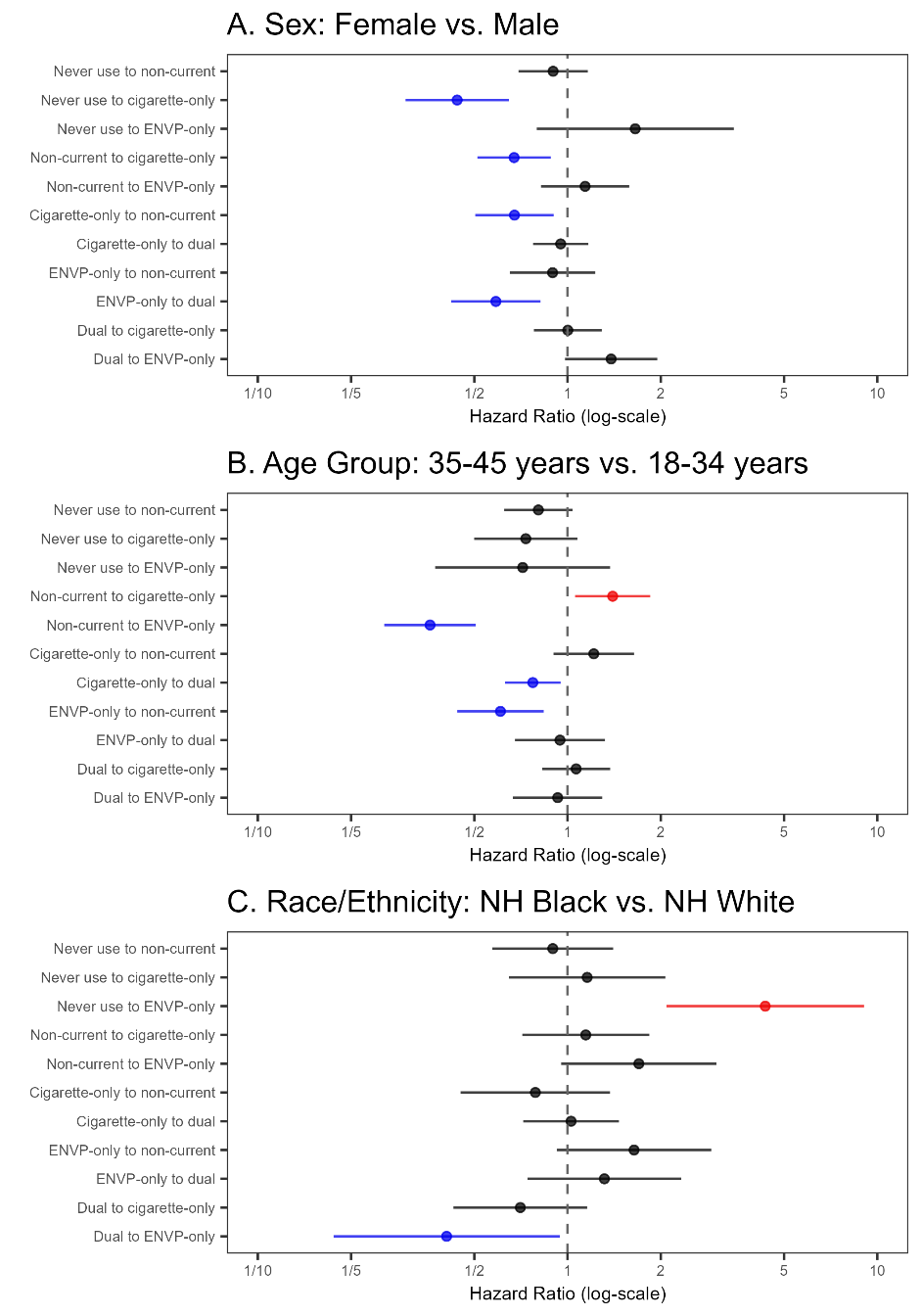

***Table S2****: One-year transition probabilities estimated across all 16 Waves of the Rutgers Omnibus Study and by period (Waves 1–5 (February 2022 to February 2023; abbreviated as 2022), 5–8 (February 2023 to January 2024; abbreviated as 2023), 8–12 (January 2024 to February 2025, abbreviated as 2024), and 12–16 (February 2025 to February 2026; abbreviated as 2025).*

| **Period** | **Transition** | **One-year transition probability** | **Lower Bound of 95% CI** | **Upper Bound of 95% CI** |
| --- | --- | --- | --- | --- |
| All Waves | Never use persistence | 82.3 | 80.7 | 83.6 |
| All Waves | Never use to non-current | 9.6 | 8.6 | 10.7 |
| All Waves | Never use to cigarette-only | 3.9 | 3.4 | 4.5 |
| All Waves | Never use to ENVP-only | 2.2 | 1.8 | 2.8 |
| All Waves | Never use to dual | 2 | 1.7 | 2.3 |
| All Waves | Non-current to never use | 0 | 0 | 0 |
| All Waves | Non-current persistence | 72.2 | 70.1 | 74.2 |
| All Waves | Non-current to cigarette-only | 11 | 9.8 | 12.4 |
| All Waves | Non-current to ENVP-only | 9.9 | 8.7 | 11.2 |
| All Waves | Non-current to dual | 7 | 6.3 | 7.7 |
| All Waves | Cigarette-only to never use | 0 | 0 | 0 |
| All Waves | Cigarette-only to non-current | 17.4 | 15.6 | 19.3 |
| All Waves | Cigarette-only persistence | 39.5 | 36.9 | 42 |
| All Waves | Cigarette-only to ENVP-only | 5.1 | 4.5 | 5.7 |
| All Waves | Cigarette-only to dual | 38 | 35.7 | 40.5 |
| All Waves | ENVP-only to never use | 0 | 0 | 0 |
| All Waves | ENVP-only to non-current | 29 | 25.7 | 32.5 |
| All Waves | ENVP-only to cigarette-only | 7.5 | 6.8 | 8.4 |
| All Waves | ENVP-only persistence | 40.1 | 36.4 | 43.7 |
| All Waves | ENVP-only to dual | 23.4 | 20.6 | 26.3 |
| All Waves | Dual to never use | 0 | 0 | 0 |
| All Waves | Dual to non-current | 7.2 | 6.5 | 7.9 |
| All Waves | Dual to cigarette-only | 22 | 20 | 24 |
| All Waves | Dual to ENVP-only | 10.1 | 8.7 | 11.6 |
| All Waves | Dual persistence | 60.8 | 58.3 | 63.2 |
| 2022 | Never use persistence | 71.8 | 67.2 | 75.1 |
| 2022 | Never use to non-current | 14.4 | 12.2 | 17.4 |
| 2022 | Never use to cigarette-only | 7.1 | 5.7 | 9 |
| 2022 | Never use to ENVP-only | 3.2 | 2.4 | 4.6 |
| 2022 | Never use to dual | 3.4 | 2.7 | 4.4 |
| 2022 | Non-current to never use | 0 | 0 | 0 |
| 2022 | Non-current persistence | 61.8 | 56.2 | 66.8 |
| 2022 | Non-current to cigarette-only | 18.5 | 15.3 | 22.4 |
| 2022 | Non-current to ENVP-only | 9.3 | 6.8 | 12.5 |
| 2022 | Non-current to dual | 10.5 | 8.7 | 12.7 |
| 2022 | Cigarette-only to never use | 0 | 0 | 0 |
| 2022 | Cigarette-only to non-current | 18.8 | 14.7 | 23.7 |
| 2022 | Cigarette-only persistence | 40.8 | 35.3 | 46.2 |
| 2022 | Cigarette-only to ENVP-only | 6.1 | 4.8 | 7.9 |
| 2022 | Cigarette-only to dual | 34.3 | 29.5 | 39.1 |
| 2022 | ENVP-only to never use | 0 | 0 | 0 |
| 2022 | ENVP-only to non-current | 28.6 | 22.8 | 36.2 |
| 2022 | ENVP-only to cigarette-only | 12.7 | 10.5 | 15 |
| 2022 | ENVP-only persistence | 31.2 | 24.1 | 38.7 |
| 2022 | ENVP-only to dual | 27.6 | 21.5 | 33.8 |
| 2022 | Dual to never use | 0 | 0 | 0 |
| 2022 | Dual to non-current | 9.7 | 7.8 | 12.3 |
| 2022 | Dual to cigarette-only | 25.6 | 21.5 | 30.2 |
| 2022 | Dual to ENVP-only | 11.8 | 9 | 15.5 |
| 2022 | Dual persistence | 52.8 | 47.3 | 58 |
| 2023 | Never use persistence | 82 | 77.4 | 85.2 |
| 2023 | Never use to non-current | 9.7 | 7.4 | 12.7 |
| 2023 | Never use to cigarette-only | 4.7 | 3.4 | 6.7 |
| 2023 | Never use to ENVP-only | 1.8 | 1.1 | 3 |
| 2023 | Never use to dual | 1.8 | 1.3 | 2.6 |
| 2023 | Non-current to never use | 0 | 0 | 0 |
| 2023 | Non-current persistence | 70 | 63.8 | 75 |
| 2023 | Non-current to cigarette-only | 14.7 | 11.4 | 18.9 |
| 2023 | Non-current to ENVP-only | 8.2 | 5.7 | 11.5 |
| 2023 | Non-current to dual | 7.1 | 5.6 | 9.1 |
| 2023 | Cigarette-only to never use | 0 | 0 | 0 |
| 2023 | Cigarette-only to non-current | 23.4 | 18.5 | 28.6 |
| 2023 | Cigarette-only persistence | 44.1 | 38.4 | 49.8 |
| 2023 | Cigarette-only to ENVP-only | 5.6 | 4.3 | 7.2 |
| 2023 | Cigarette-only to dual | 26.9 | 22.2 | 31.8 |
| 2023 | ENVP-only to never use | 0 | 0 | 0 |
| 2023 | ENVP-only to non-current | 35.1 | 27.6 | 42.9 |
| 2023 | ENVP-only to cigarette-only | 14.9 | 12 | 18.6 |
| 2023 | ENVP-only persistence | 27.2 | 20 | 35.2 |
| 2023 | ENVP-only to dual | 22.7 | 17.6 | 28.9 |
| 2023 | Dual to never use | 0 | 0 | 0 |
| 2023 | Dual to non-current | 14.1 | 11.5 | 17.2 |
| 2023 | Dual to cigarette-only | 33.8 | 28.9 | 39.1 |
| 2023 | Dual to ENVP-only | 10.7 | 7.9 | 14 |
| 2023 | Dual persistence | 41.4 | 35.6 | 46.9 |
| 2024 | Never use persistence | 78.3 | 73.4 | 81.8 |
| 2024 | Never use to non-current | 11.9 | 9.3 | 15.5 |
| 2024 | Never use to cigarette-only | 4.2 | 3 | 5.9 |
| 2024 | Never use to ENVP-only | 2.8 | 1.8 | 4.7 |
| 2024 | Never use to dual | 2.8 | 2 | 3.9 |
| 2024 | Non-current to never use | 0 | 0 | 0 |
| 2024 | Non-current persistence | 72.3 | 66.4 | 77.2 |
| 2024 | Non-current to cigarette-only | 10.1 | 7.4 | 13.3 |
| 2024 | Non-current to ENVP-only | 9.6 | 6.7 | 13.5 |
| 2024 | Non-current to dual | 8 | 6.1 | 10.3 |
| 2024 | Cigarette-only to never use | 0 | 0 | 0 |
| 2024 | Cigarette-only to non-current | 15 | 11.1 | 20.9 |
| 2024 | Cigarette-only persistence | 31.7 | 25.9 | 37.6 |
| 2024 | Cigarette-only to ENVP-only | 4.7 | 3.4 | 6.6 |
| 2024 | Cigarette-only to dual | 48.6 | 42.4 | 54.4 |
| 2024 | ENVP-only to never use | 0 | 0 | 0 |
| 2024 | ENVP-only to non-current | 28.2 | 21.3 | 37.8 |
| 2024 | ENVP-only to cigarette-only | 5.1 | 3.9 | 7 |
| 2024 | ENVP-only persistence | 44.3 | 33.8 | 53.2 |
| 2024 | ENVP-only to dual | 22.4 | 15.3 | 31.3 |
| 2024 | Dual to never use | 0 | 0 | 0 |
| 2024 | Dual to non-current | 4.8 | 3.6 | 6.8 |
| 2024 | Dual to cigarette-only | 14.4 | 10.7 | 18.8 |
| 2024 | Dual to ENVP-only | 8.5 | 5.4 | 12.5 |
| 2024 | Dual persistence | 72.3 | 65.6 | 77.6 |
| 2025 | Never use persistence | 74.3 | 67.8 | 78.4 |
| 2025 | Never use to non-current | 13.2 | 10.5 | 17.1 |
| 2025 | Never use to cigarette-only | 5.1 | 3.6 | 7.4 |
| 2025 | Never use to ENVP-only | 3.8 | 2.6 | 6.2 |
| 2025 | Never use to dual | 3.6 | 2.8 | 5.1 |
| 2025 | Non-current to never use | 0 | 0 | 0 |
| 2025 | Non-current persistence | 65.6 | 59.7 | 70.9 |
| 2025 | Non-current to cigarette-only | 10.5 | 7.8 | 13.8 |
| 2025 | Non-current to ENVP-only | 13.5 | 10.2 | 17.4 |
| 2025 | Non-current to dual | 10.5 | 8.3 | 13.2 |
| 2025 | Cigarette-only to never use | 0 | 0 | 0 |
| 2025 | Cigarette-only to non-current | 21.1 | 15.8 | 28 |
| 2025 | Cigarette-only persistence | 30.5 | 23.7 | 37 |
| 2025 | Cigarette-only to ENVP-only | 7.9 | 6.2 | 10.3 |
| 2025 | Cigarette-only to dual | 40.5 | 33.6 | 46.9 |
| 2025 | ENVP-only to never use | 0 | 0 | 0 |
| 2025 | ENVP-only to non-current | 35 | 27.9 | 42.3 |
| 2025 | ENVP-only to cigarette-only | 8.8 | 6.8 | 11.3 |
| 2025 | ENVP-only persistence | 28.6 | 21.1 | 36.5 |
| 2025 | ENVP-only to dual | 27.6 | 20.9 | 35.5 |
| 2025 | Dual to never use | 0 | 0 | 0 |
| 2025 | Dual to non-current | 10.7 | 8.5 | 14.1 |
| 2025 | Dual to cigarette-only | 18.3 | 14.2 | 22.9 |
| 2025 | Dual to ENVP-only | 12.2 | 9 | 16.6 |
| 2025 | Dual persistence | 58.8 | 52.5 | 64.7 |

***Table S3****. All unadjusted and adjusted transition hazard ratios estimated across all 16 waves of the Rutgers Omnibus Study (February 2022 to February 2025) by covariate (sex, age group, and race & ethnicity) with confidence intervals.*

| **Model** | **Covariate** | **Hazard Ratio** | **Lower bound of 95% CI** | **Upper bound of 95% CI** |
| --- | --- | --- | --- | --- |
| Unadjusted | Female | 0.90 | 0.69 | 1.16 |
| Unadjusted | Female | 0.44 | 0.30 | 0.65 |
| Unadjusted | Female | 1.65 | 0.80 | 3.44 |
| Unadjusted | Female | 0.67 | 0.51 | 0.88 |
| Unadjusted | Female | 1.14 | 0.82 | 1.58 |
| Unadjusted | Female | 0.67 | 0.50 | 0.90 |
| Unadjusted | Female | 0.95 | 0.77 | 1.17 |
| Unadjusted | Female | 0.89 | 0.65 | 1.23 |
| Unadjusted | Female | 0.59 | 0.42 | 0.82 |
| Unadjusted | Female | 1.00 | 0.78 | 1.29 |
| Unadjusted | Female | 1.38 | 0.98 | 1.95 |
| Unadjusted | 35-45 | 0.81 | 0.62 | 1.04 |
| Unadjusted | 35-45 | 0.73 | 0.50 | 1.08 |
| Unadjusted | 35-45 | 0.72 | 0.37 | 1.37 |
| Unadjusted | 35-45 | 1.40 | 1.06 | 1.85 |
| Unadjusted | 35-45 | 0.36 | 0.26 | 0.51 |
| Unadjusted | 35-45 | 1.22 | 0.90 | 1.64 |
| Unadjusted | 35-45 | 0.77 | 0.63 | 0.95 |
| Unadjusted | 35-45 | 0.61 | 0.44 | 0.84 |
| Unadjusted | 35-45 | 0.95 | 0.68 | 1.32 |
| Unadjusted | 35-45 | 1.07 | 0.83 | 1.37 |
| Unadjusted | 35-45 | 0.93 | 0.67 | 1.29 |
| Unadjusted | Black | 0.90 | 0.57 | 1.40 |
| Unadjusted | Black | 1.16 | 0.65 | 2.07 |
| Unadjusted | Black | 4.35 | 2.09 | 9.05 |
| Unadjusted | Black | 1.15 | 0.71 | 1.84 |
| Unadjusted | Black | 1.70 | 0.95 | 3.03 |
| Unadjusted | Black | 0.79 | 0.45 | 1.37 |
| Unadjusted | Black | 1.03 | 0.72 | 1.47 |
| Unadjusted | Black | 1.64 | 0.92 | 2.91 |
| Unadjusted | Black | 1.31 | 0.74 | 2.33 |
| Unadjusted | Black | 0.70 | 0.43 | 1.16 |
| Unadjusted | Black | 0.41 | 0.18 | 0.94 |
| Adjusted | Female | 0.95 | 0.70 | 1.28 |
| Adjusted | Female | 0.44 | 0.29 | 0.66 |
| Adjusted | Female | 1.29 | 0.58 | 2.86 |
| Adjusted | Female | 0.65 | 0.48 | 0.88 |
| Adjusted | Female | 1.07 | 0.71 | 1.63 |
| Adjusted | Female | 0.59 | 0.42 | 0.81 |
| Adjusted | Female | 0.98 | 0.78 | 1.24 |
| Adjusted | Female | 0.90 | 0.61 | 1.33 |
| Adjusted | Female | 0.54 | 0.37 | 0.80 |
| Adjusted | Female | 0.87 | 0.66 | 1.16 |
| Adjusted | Female | 1.37 | 0.94 | 1.99 |
| Adjusted | 35-45 | 0.84 | 0.63 | 1.12 |
| Adjusted | 35-45 | 0.64 | 0.42 | 0.98 |
| Adjusted | 35-45 | 0.65 | 0.31 | 1.35 |
| Adjusted | 35-45 | 1.39 | 1.02 | 1.90 |
| Adjusted | 35-45 | 0.34 | 0.23 | 0.51 |
| Adjusted | 35-45 | 1.45 | 1.03 | 2.04 |
| Adjusted | 35-45 | 0.74 | 0.59 | 0.93 |
| Adjusted | 35-45 | 0.59 | 0.40 | 0.85 |
| Adjusted | 35-45 | 1.11 | 0.76 | 1.63 |
| Adjusted | 35-45 | 1.05 | 0.79 | 1.40 |
| Adjusted | 35-45 | 0.98 | 0.68 | 1.39 |
| Adjusted | Black | 0.92 | 0.58 | 1.45 |
| Adjusted | Black | 1.11 | 0.62 | 1.99 |
| Adjusted | Black | 4.39 | 2.09 | 9.25 |
| Adjusted | Black | 1.25 | 0.78 | 2.01 |
| Adjusted | Black | 1.74 | 0.95 | 3.19 |
| Adjusted | Black | 0.69 | 0.38 | 1.24 |
| Adjusted | Black | 1.02 | 0.72 | 1.46 |
| Adjusted | Black | 1.74 | 0.95 | 3.17 |
| Adjusted | Black | 1.31 | 0.73 | 2.37 |
| Adjusted | Black | 0.68 | 0.41 | 1.14 |
| Adjusted | Black | 0.47 | 0.21 | 1.04 |
